# Protocol for the Development and Prospective Evaluation of ASHA Assist India: An AI-Assisted Mobile Platform for Community-Based Stroke Prevention in Rural India

**DOI:** 10.64898/2026.08.10.26360065

**Authors:** Karthik Samdeep Nayak, Abhay S Nirgude, Ranajit Das

**Affiliations:** Centre for Integrative Research in Clinical Genetics and Metabolic Medicine, Yenepoya Medical College, Yenepoya (Deemed to be University), Mangalore, Karnataka, India; Department of Community Medicine, Yenepoya Medical College, Yenepoya (Deemed to be University), Mangalore, Karnataka, India; Centre for Systems Biology and Molecular Medicine, Yenepoya Research Centre, Yenepoya (Deemed to be University), Mangalore, Karnataka, India

**Keywords:** Stroke prevention, Mobile health (mHealth), Artificial intelligence, Community health workers, Accredited Social Health Activists (ASHAs), Digital health, Implementation science, Primary healthcare, Clinical decision support, Health informatics, Rural health, Ayushman Bharat Digital Mission (ABDM), Protocol

## Abstract

**Background:** Stroke remains one of the leading causes of mortality and long-term disability worldwide, with low- and middle-income countries bearing a disproportionate share of the global disease burden. In India, delays in risk identification, fragmented referral pathways, and limited continuity of preventive care present significant challenges, particularly in rural communities. As a frontline health worker Accredited Social Health Activists (ASHAs) are strategically positioned to support community-based stroke prevention; however, existing workflows are frequently constrained by multi-tasking, predominantly paper-based documentation and fragmented digital systems. Advances in mobile health, artificial intelligence along with digital health ecosystem provided by Ayushman Bharat Digital Mission (ABDM) provide an opportunity to strengthen community healthcare through integrated digital platforms.

**Objective:** This protocol describes the design, system architecture, and prospective evaluation framework of ASHA Assist India, an integrated AI-assisted mobile health platform intended to support community-based stroke prevention by connecting citizens, ASHA workers, Primary Health Centres (PHCs), and higher levels of healthcare facilities within a unified digital ecosystem.

**Methods:** ASHA Assist India has been designed as a modular, cloud-based digital health platform supporting standardized data collection, longitudinal health monitoring, referral management, and AI-assisted clinical decision support. The proposed system comprises four user-facing applications corresponding to citizens, ASHA workers, PHCs, and referral hospitals, integrated through a centralized backend providing authentication, secure data management, interoperability, analytics, and notification services. The AI framework includes three planned analytical modules: (i) population-level stroke risk stratification, (ii) longitudinal stroke risk prediction, and (iii) acute stroke symptom recognition. A prospective implementation study is planned to evaluate platform usability, feasibility, workflow integration, implementation outcomes, and operational performance within routine community healthcare settings. Future validation of the AI modules will be conducted using prospectively collected longitudinal datasets.

**Expected Impact:** The proposed platform aims to strengthen community-based stroke prevention by improving digital workflow integration, facilitating coordinated referral pathways, and supporting longitudinal monitoring through the existing healthcare providers at health and wellness centres like ASHA, Community Health Officers (CHOs), ANM, etc. Beyond stroke prevention, the modular architecture is intended to provide a scalable framework for future digital health programmes addressing multiple non-communicable diseases within primary healthcare systems. Publication of this protocol establishes a transparent implementation and evaluation framework that may guide future research, digital health innovation, and implementation science in resource-constrained settings.

**Protocol Status and Scope:** This manuscript presents the protocol and technical design of the ASHA Assist India platform. It does not report findings from pilot deployment, implementation studies, or clinical validation. The manuscript describes the proposed system architecture, functional components, implementation strategy, governance framework, and prospective evaluation plan. Software development, pilot implementation, ethics approval, and validation of the AI-assisted decision-support modules will be undertaken in subsequent phases of the project.

## Introduction

Stroke remains one of the leading causes of death and long-term disability worldwide, accounting for an estimated 7.3 million deaths and more than 160 million disability-adjusted life years (DALYs) in 2021. Although mortality rates have declined in many high-income countries owing to improvements in prevention and acute stroke care, the overall global burden of stroke continues to rise because of population ageing, increasing prevalence of hypertension, diabetes mellitus, obesity, and other modifiable vascular risk factors. Importantly, nearly 90% of stroke-related deaths and disability occur in low- and middle-income countries, where access to preventive healthcare and specialist neurological services remains limited (1,2).

India contributes substantially to the global stroke burden, with recent estimates suggesting approximately 1.8 million new stroke cases annually. Stroke occurs at a relatively younger age in the Indian population compared with many high-income countries and remains a major contributor to premature mortality, disability, and healthcare expenditure. Delayed recognition of stroke symptoms, inadequate screening for vascular risk factors, fragmented referral pathways, and unequal access to specialist care continue to limit timely intervention, particularly in rural and underserved regions (1,3,4). Because the effectiveness of acute stroke treatment is highly time-dependent, strengthening community-level prevention and improving early identification of individuals at increased risk are essential components of national stroke control strategies.

India possesses one of the world’s largest community health worker programmes through its network of Accredited Social Health Activists (ASHAs). Established under the National Health Mission, ASHAs serve as the primary link between rural communities and the formal healthcare system by promoting health awareness, conducting household visits, facilitating disease prevention activities, and supporting referrals to higher levels of care. Their close engagement with households positions them uniquely to support community-based screening and longitudinal monitoring of individuals at risk of chronic diseases, including stroke. However, despite increasing digitalization of healthcare services, routine stroke prevention activities remain largely dependent on paper-based documentation or isolated digital applications that are not fully integrated into community workflows or referral systems.

Recent advances in mobile health (mHealth), interoperable health information systems, and artificial intelligence (AI) provide opportunities to strengthen community-based healthcare delivery. Mobile applications enable standardized data collection, longitudinal monitoring, secure communication between healthcare providers, and timely referral of high-risk individuals. Artificial intelligence has further demonstrated potential for supporting clinical decision-making through automated risk stratification and decision-support tools. Nevertheless, successful implementation of digital health interventions depends not only on analytical performance but also on usability, workflow integration, interoperability, offline functionality, data governance, and acceptance by frontline healthcare workers. The World Health Organization’s Global Strategy on Digital Health emphasizes that digital technologies should strengthen existing health systems rather than create parallel service delivery models (5).

Recognizing these implementation challenges, we propose ASHA Assist India, an integrated mobile health platform designed to support community-based stroke prevention through the existing ASHA network. Rather than functioning solely as an artificial intelligence application, the proposed platform is intended to provide an integrated digital ecosystem connecting citizens, ASHA workers, Primary Health Centres (PHCs), and referral hospitals. The platform is designed to support standardized data collection, longitudinal follow-up, referral management, and clinical decision support while maintaining compatibility with India’s evolving digital health ecosystem, including the Ayushman Bharat Digital Mission (ABDM).

This manuscript presents the protocol and technical design of the ASHA Assist India platform. Specifically, it describes the proposed system architecture, user workflows, software components, implementation strategy, and prospective evaluation framework that will guide future pilot deployment and assessment of feasibility, usability, workflow integration, and implementation outcomes. By publishing the protocol before large-scale implementation, we aim to provide a transparent framework for the development and evaluation of community-based digital health interventions for stroke prevention in rural India.

## 2. Study Objectives and Overall Protocol Design

### 2.1 Study Objectives

#### Primary Objective

The primary objective of the ASHA Assist India project is to design, implement, and prospectively evaluate an integrated mobile health platform that supports community-based stroke prevention through India’s existing Accredited Social Health Activist (ASHA) network. The platform is intended to facilitate standardized stroke risk assessment, longitudinal health monitoring, clinical decision support, referral management, and communication across multiple levels of the rural healthcare system.

#### Secondary Objectives

The secondary objectives of the project are:

- To develop a unified mobile platform supporting four user groups, namely citizens, ASHA workers, Primary Health Centres (PHCs), and referral hospitals.
- To integrate structured health data collection, longitudinal monitoring, and referral workflows within a single interoperable digital ecosystem.
- To incorporate artificial intelligence–based decision support modules for population-level risk stratification and future longitudinal stroke prediction.
- To evaluate the usability, feasibility, and acceptability of the platform among ASHA workers and healthcare professionals during a prospective pilot study.
- To assess workflow integration, referral efficiency, and implementation outcomes following field deployment.

### 2.2 Overall Study Design

ASHA Assist India is designed as a prospective digital health implementation project consisting of four sequential phases (Figure 1). The present manuscript describes the protocol and technical design of the proposed platform prior to pilot implementation.

**Figure 1.**
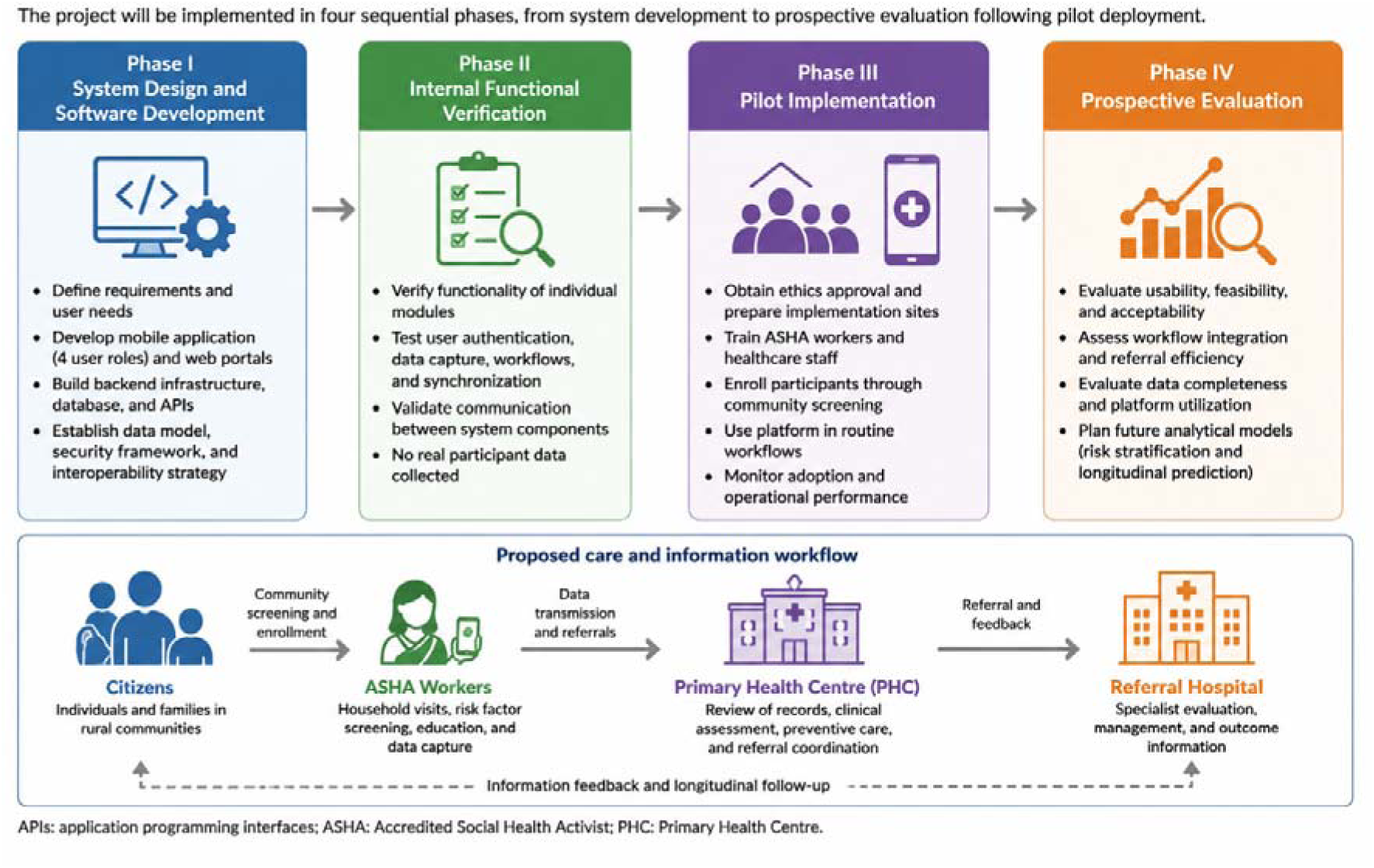
Overall protocol design of the ASHA Assist India project.

#### Phase I – System Design and Software Development

The initial phase involves the development of the mobile application, backend infrastructure, user interfaces, and system architecture supporting four user roles within the rural healthcare system. During this phase, the data model, user workflows, security framework, and interoperability strategy are established.

#### Phase II – Internal Functional Verification

Following software development, functional verification will be performed to ensure that individual software components operate according to the intended design specifications. This phase includes verification of user authentication, data capture, referral workflows, synchronization mechanisms, and communication between system components. No real participant data will be collected during this phase.

#### Phase III – Pilot Implementation

Following Institutional Ethics Committee approval, the platform will undergo prospective pilot deployment within selected rural communities. The pilot study will evaluate the feasibility of integrating the platform into routine ASHA workflows and will assess user adoption, operational performance, and workflow integration.

#### Phase IV – Prospective Evaluation

The final phase will evaluate implementation outcomes using predefined process and usability measures. Planned outcomes include user acceptability, workflow efficiency, data completeness, referral patterns, platform utilization, and operational feasibility. Future analytical components, including longitudinal stroke prediction and real-time symptom assessment, will be developed following acquisition of appropriate longitudinal datasets.

### 2.3 Overall Workflow

The proposed workflow follows the existing hierarchy of India’s rural healthcare delivery system. Citizens are enrolled through ASHA workers during routine community visits. Information collected during household screening is securely transmitted to the assigned Primary Health Centre, where healthcare professionals review participant records, coordinate preventive interventions, and initiate referrals when appropriate. Referral hospitals provide specialist evaluation and communicate clinical outcomes back through the same digital pathway, enabling continuity of care and longitudinal follow-up.

Artificial intelligence–based analytical modules are designed to function as clinical decision-support tools within this workflow rather than replacing clinical judgement. Risk estimates generated by these modules are intended to assist healthcare workers in prioritizing preventive interventions and referrals while preserving clinician oversight throughout the decision-making process.

## 3. System Architecture

### 3.1 Design Philosophy

The ASHA Assist India platform is designed as an integrated digital health ecosystem that supports community-based stroke prevention across multiple levels of the rural healthcare system. The proposed architecture follows a user-centered, modular design philosophy, enabling different categories of users to perform role-specific tasks while maintaining secure data exchange and continuity of care throughout the referral pathway.

Unlike standalone mobile applications that focus exclusively on risk prediction or data collection, the proposed platform is intended to support the complete continuum of preventive care, beginning with community-based screening and extending through referral, clinical assessment, follow-up, and longitudinal monitoring. The architecture therefore emphasizes interoperability, scalability, usability, and compatibility with existing healthcare workflows rather than functioning solely as an artificial intelligence application.

To achieve these objectives, the platform adopts a multi-tier architecture consisting of four interacting user domains Citizens, Accredited Social Health Activists (ASHAs), Primary Health Centres (PHCs), and Referral Hospitals both public and private —supported by a centralized cloud-based infrastructure responsible for authentication, data management, clinical decision support, and secure communication. This architecture is designed to ensure that each stakeholder accesses information appropriate to their responsibilities while enabling seamless information flow across different levels of healthcare delivery.

### 3.2 Overall System Architecture

The proposed architecture comprises four principal layers that collectively support data acquisition, information management, clinical decision support, and healthcare delivery (Figure 2).

**Figure 2.**
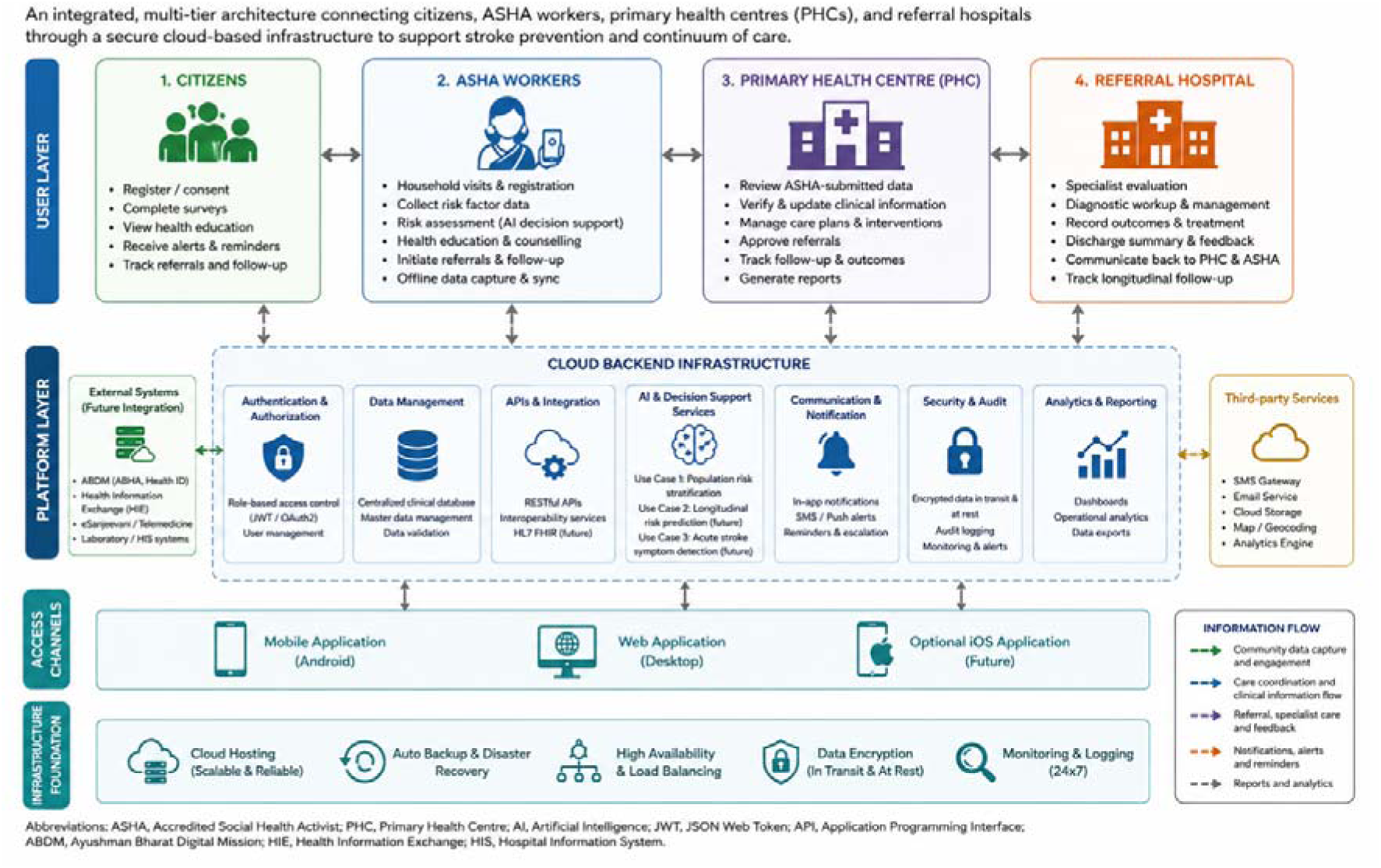
System Architecture of ASHA Assist India Platform.

**Figure 3.**
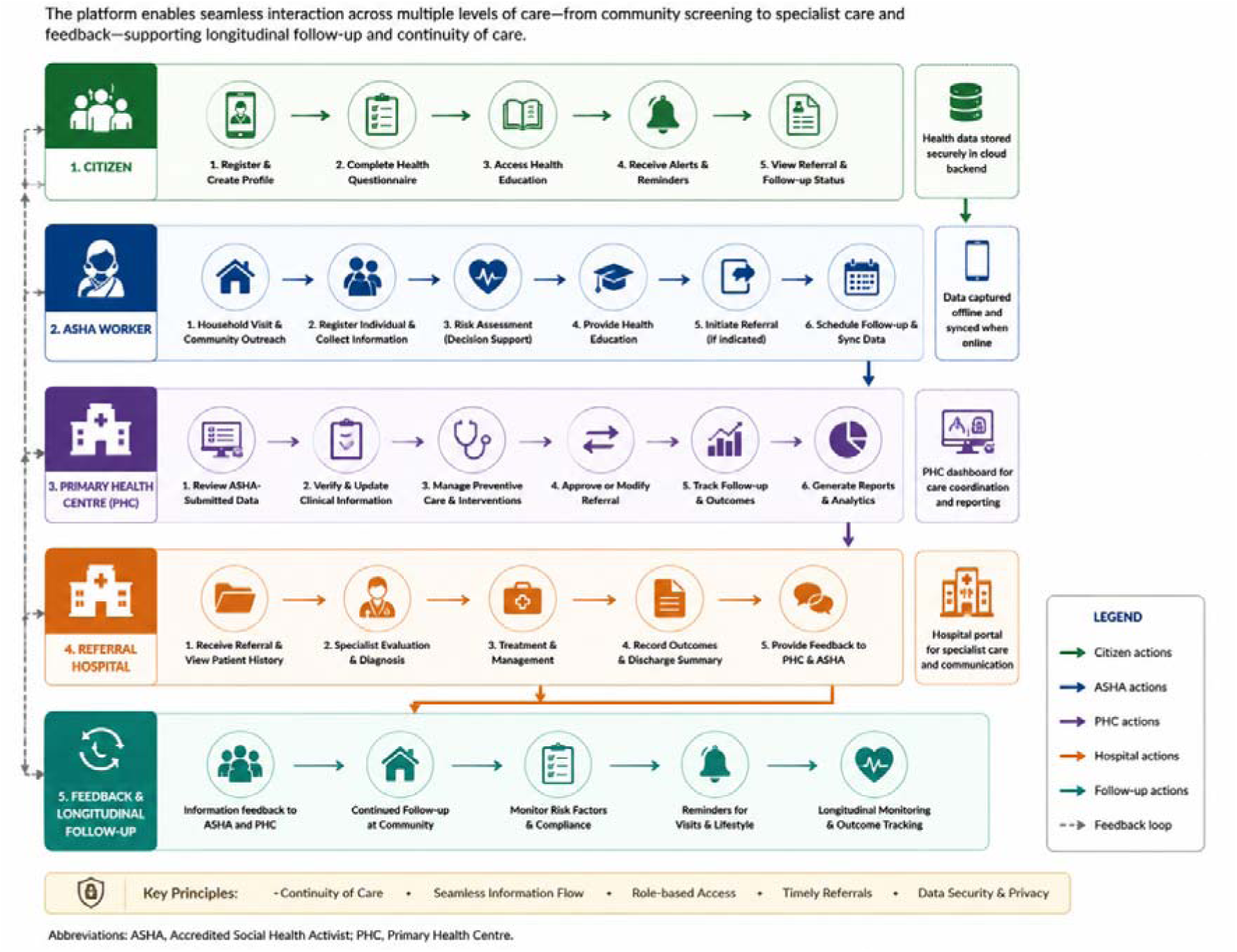
User Journey and Functional Workflow in ASHA Assist India Platform.

#### Community Layer

The community layer represents the first point of interaction with the healthcare system and includes citizens participating in community-based screening programmes. Demographic information, lifestyle characteristics, medical history, and other relevant health indicators are collected during household visits or community outreach activities performed by ASHA workers. The platform is designed to facilitate standardized digital data capture while minimizing documentation burden by integrating with existing digital solutions and improving data completeness.

#### Community Health Worker Layer

ASHA workers constitute the operational interface between rural communities and the formal healthcare system. Through a dedicated mobile application, ASHAs are intended to conduct participant registration, collect health information, perform structured risk assessments, deliver health education, schedule follow-up visits, and initiate referrals when indicated. The application is designed to support offline data capture with subsequent synchronization when network connectivity becomes available, recognizing the challenges of intermittent internet access in many rural settings. Application will assist ASHA in clinical decision support, appropriate referral and continuum of care. Significant time ASHA or any frontline workers needs to spend on preparing the report and timely submission of report in various formats as per the different program guidelines. This platform will reduce that burden and ease the process and facilitate availability of reports in realtime.

#### Primary Healthcare Layer

Primary Health Centres function as the first level of clinical review within the proposed platform. Healthcare professionals at PHCs are expected to review participant records submitted by ASHA workers, verify screening information, coordinate preventive interventions, and determine whether referral to higher-level facilities is warranted. The platform is designed to provide centralized access to participant records while maintaining role-based access control and audit trails.

#### Referral Hospital Layer

Referral hospitals represent the highest level of care within the proposed workflow. Following referral, specialists are expected to review participant information, document diagnostic findings, initiate appropriate management, and communicate clinical outcomes through the platform. Feedback generated during specialist evaluation is intended to become available to PHCs and ASHA workers, supporting continuity of care and longitudinal follow-up after hospital discharge.

### 3.3 Backend Infrastructure

The proposed backend infrastructure is designed around a secure cloud-based architecture that manages user authentication, data storage, application programming interfaces (APIs), and communication among system components. The backend serves as the central coordination layer, enabling synchronized information exchange across multiple user interfaces while maintaining data integrity and security.

Key architectural components include:

- Secure user authentication and role-based authorization.
- Centralized clinical database.
- RESTful application programming interfaces for data exchange.
- Encrypted communication between mobile and web applications.
- Audit logging for traceability.
- Offline synchronization mechanisms to accommodate limited network connectivity.
- Modular analytical services supporting future artificial intelligence integration.
- The modular design is intended to facilitate future expansion without requiring substantial redesign of the overall platform architecture.

### 3.4 Design Principles

The architecture has been developed according to several guiding design principles:

- Modularity: Individual software components are designed to function independently, facilitating future development and maintenance.
- Scalability: The architecture is intended to support expansion from pilot implementation to larger regional or national deployments.
- Interoperability: Standardized APIs are proposed to facilitate integration with existing digital health ecosystems and future national health information platforms.
- Security and Privacy: User authentication, encrypted communication, role-based permissions, and audit logging are incorporated into the design to support secure management of health information.
- Offline Capability: Recognizing connectivity challenges in rural India, the platform is designed to permit offline data collection with automatic synchronization once internet access becomes available.
- Human-Centred Design: Clinical decision support is intended to assist, rather than replace, healthcare workers. Final clinical decisions remain the responsibility of qualified healthcare professionals.

## 4. Functional Components of the ASHA Assist India Platform

### 4.1 Overview

The ASHA Assist India platform is designed as a modular digital health ecosystem comprising multiple interoperable components that collectively support community-based stroke prevention, longitudinal health monitoring, referral management, and clinical decision support. Each module has been developed to address a specific stage of the healthcare pathway while maintaining seamless communication with the centralized backend infrastructure. This modular architecture is intended to facilitate future expansion and integration with additional healthcare services without requiring substantial redesign of the underlying system.

The platform is organized into four principal functional modules corresponding to the primary user groups within the proposed healthcare workflow: (i) Citizen Application, (ii) ASHA Worker Application, (iii) Primary Health Centre Dashboard, and (iv) Referral Hospital Portal. Supporting these user-facing applications is a centralized cloud backend responsible for authentication, data management, interoperability, notifications, analytics, and future AI-enabled decision support.

### 4.2 Citizen Application

The Citizen Application is designed to serve as the public-facing component of the platform, enabling individuals to participate actively in community-based stroke prevention. The application is intended to facilitate participant registration, electronic consent, completion of health questionnaires, and access to personalized educational resources related to stroke prevention and healthy lifestyle practices.

Planned features of the Citizen Application include:

- Secure participant registration and authentication.
- Digital informed consent.
- Health and lifestyle questionnaires.
- Stroke prevention educational resources.
- Appointment reminders and notifications.
- Referral tracking and follow-up status.
- Access to personalized health records.

The application is designed with a simplified user interface to maximize accessibility for individuals with varying levels of digital literacy.

### 4.3 ASHA Worker Application

The ASHA Worker Application represents the primary operational interface of the proposed platform and is intended to support routine community health activities. During household visits, ASHA workers will use the application to register participants, collect demographic and clinical information, document lifestyle risk factors, provide health education, and initiate referrals when appropriate.

The application is designed to support:

- Participant registration.
- Household-based screening.
- Structured data collection.
- Risk factor documentation.
- Health analytics
- Clinical decision-support prompts.
- Referral generation.
- Follow-up scheduling.
- Offline data collection with automatic synchronization when network connectivity becomes available.

The interface is designed to minimize documentation burden while preserving standardized data collection across different field settings.

### 4.4 Primary Health Centre Dashboard

Healthcare professionals at Primary Health Centres (PHCs) will access participant information through a secure web-based dashboard. The dashboard is intended to facilitate review of screening records submitted by ASHA workers, verification of clinical information, management of preventive interventions, and coordination of referrals to higher-level healthcare facilities.

Planned dashboard functionality includes:

- Review of participant records.
- Verification and updating of clinical information.
- Monitoring of referral status.
- Longitudinal Individual/Population health trajectory
- Population health summaries.
- Follow-up management.
- Clinical reporting.

Role-based access controls are intended to ensure that users can access only information relevant to their professional responsibilities.

### 4.5 Referral Hospital Portal

The Referral Hospital Portal is designed to support specialist assessment and communication following referral from Primary Health Centres. Healthcare providers will be able to review participant histories, document diagnostic findings, record management decisions, and communicate clinical outcomes back to referring facilities.

Planned capabilities include:

- Specialist case review.
- Diagnostic documentation.
- Treatment recording.
- Discharge summaries.
- Communication with PHCs.
- Longitudinal follow-up documentation.

This module is intended to strengthen continuity of care by enabling bidirectional information exchange between secondary and primary healthcare services.

### 4.6 Shared Platform Services

Several supporting services are designed to operate across all user modules. These include:

- User authentication and authorization.
- Consent management system?
- Role-based access control.
- Centralized clinical database.
- Secure application programming interfaces (APIs).
- Notification services.
- Audit logging.
- Data encryption.
- Analytics and reporting.
- Future interoperability with national digital health infrastructure.

These shared services are intended to provide a common technological foundation for all platform components while ensuring security, scalability, and maintainability.

## 5. Artificial Intelligence Framework and Decision Support Modules

### 5.1 Overview

Artificial intelligence (AI) within the ASHA Assist India platform is conceived as a clinical decision-support component rather than an autonomous diagnostic system. The proposed AI framework is intended to complement existing healthcare workflows by assisting community health workers and clinicians in identifying individuals who may benefit from preventive interventions, further clinical evaluation, or specialist referral. All AI-generated recommendations are designed to support—not replace—the clinical judgement of qualified healthcare professionals.

The AI framework adopts a modular architecture, enabling independent development, validation, and deployment of analytical models according to data availability and clinical requirements. This approach allows individual models to be updated or replaced without affecting the overall functionality of the platform.

### 5.2 Planned AI Modules

Three analytical modules are proposed as part of the ASHA Assist India platform (Figure 4).

**Figure 4.**
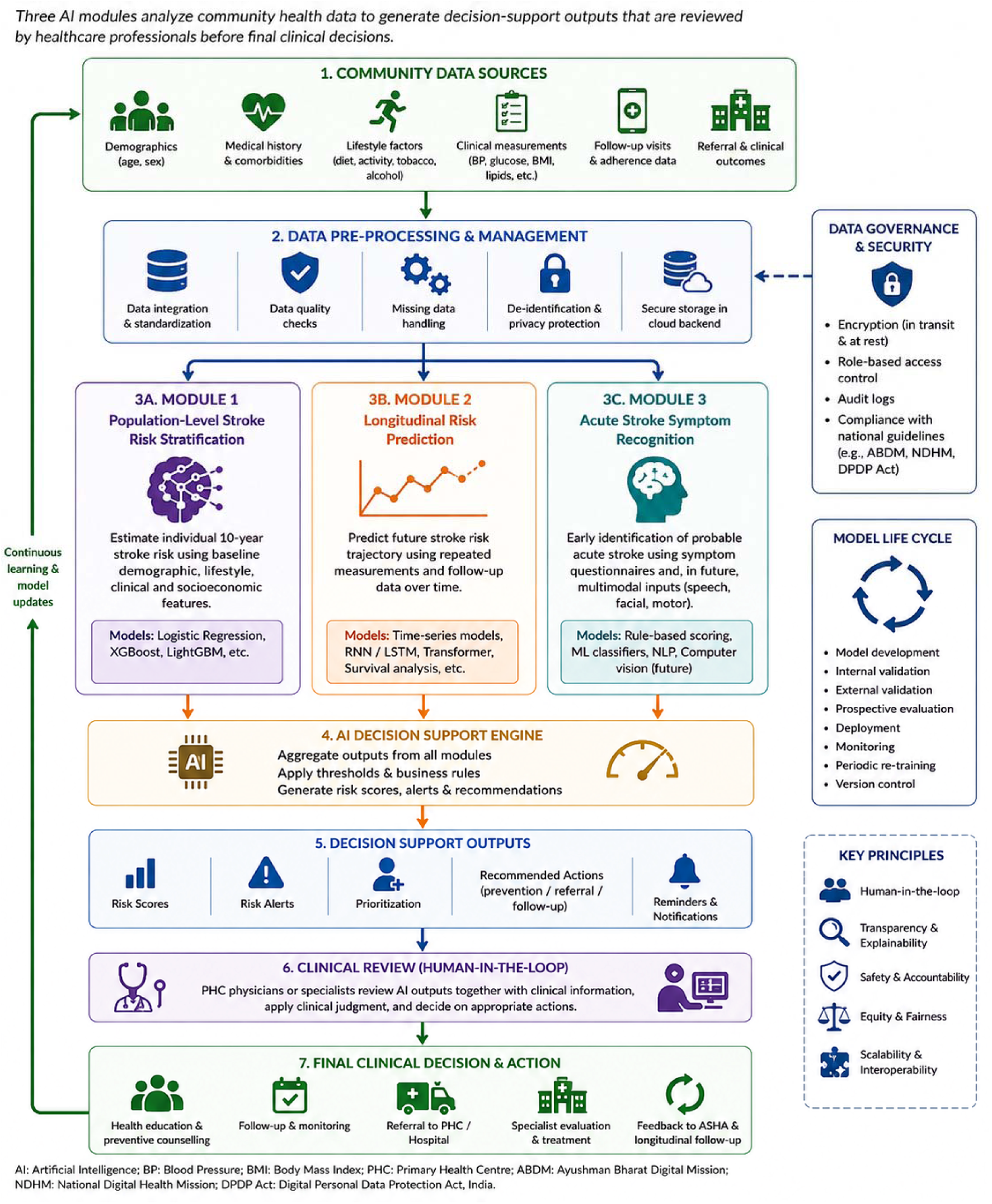
Proposed Al Framework and Decision Support Modules in ASHA Assist India Platform.

**Figure 5.**
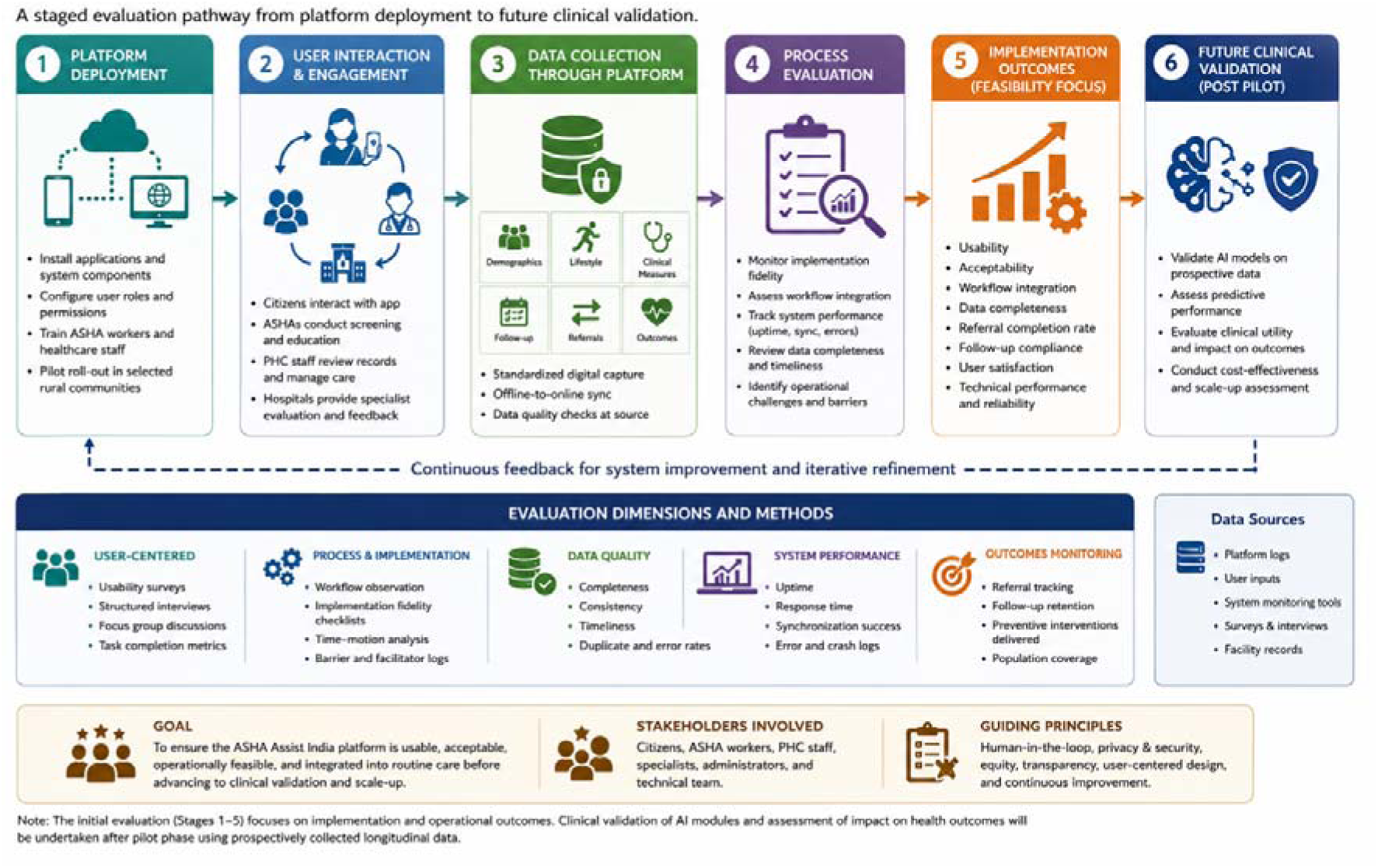
Prospective Evaluation Framework for ASHA Assist India Platform.

**Figure 6.**
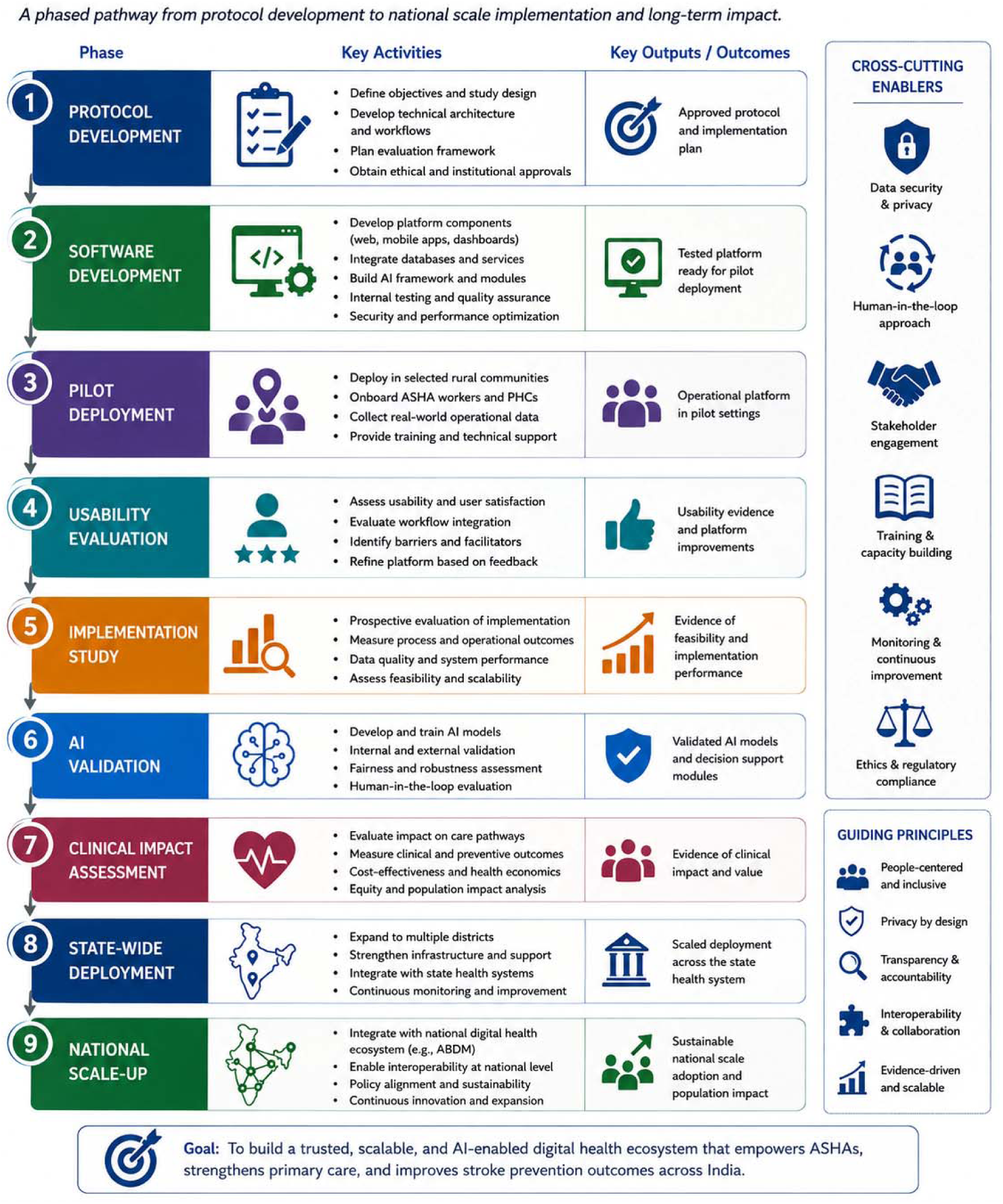
Roadmap for Progressive Development and Scale-up of ASHA Assist India.

#### Module 1: Population-Level Stroke Risk Stratification

The first module is designed to estimate an individual’s probability of stroke using routinely collected demographic, lifestyle, socioeconomic, and clinical variables. This module is intended to support community-based screening by identifying individuals who may benefit from preventive counselling or referral for further clinical evaluation.

The analytical framework will be based on established statistical and machine learning methodologies, with model development, calibration, and external validation performed using appropriately curated datasets. An initial proof-of-concept model has been developed separately using publicly available epidemiological data and is reported independently; future versions will be retrained using prospectively collected Indian population data to improve clinical relevance and generalizability.

#### Module 2: Longitudinal Risk Prediction

The second module is designed to analyse repeated observations collected during routine follow-up visits to estimate changes in stroke risk over time. Rather than relying on a single assessment, this module will incorporate longitudinal health information to monitor temporal trends in modifiable risk factors, treatment adherence, and disease progression.

Future analytical approaches may include time-series modelling, recurrent neural networks, transformer-based architectures, or survival analysis methods, depending on the characteristics and volume of longitudinal data collected during prospective deployment.

#### Module 3: Acute Stroke Symptom Recognition

The third module is intended to support rapid recognition of acute stroke symptoms within the community. This component will evaluate participant-reported symptoms using structured clinical questionnaires based on established stroke recognition tools and may, in future versions, incorporate multimodal inputs such as speech analysis, facial asymmetry assessment, or motor function evaluation where clinically appropriate and ethically approved.

This module is intended solely as a decision-support aid to facilitate timely referral and does not constitute a diagnostic tool.

### 5.3 Human-in-the-Loop Decision Support

All AI outputs generated by the platform are intended to be reviewed by healthcare professionals before clinical action is taken. Risk estimates, alerts, and recommendations are designed to assist prioritization and workflow management rather than making autonomous healthcare decisions.

The proposed framework follows a human-in-the-loop paradigm in which:

- ASHA workers perform data collection and initial screening.
- AI models generate risk estimates and decision-support recommendations.
- Primary healthcare professionals review AI outputs alongside clinical information.
- Final referral and treatment decisions remain the responsibility of qualified clinicians.

This design is intended to promote transparency, accountability, and safe integration of AI into routine healthcare practice.

### 5.4 Model Governance and Future Validation

Because the analytical modules are intended for use in healthcare settings, all models will undergo prospective validation before clinical deployment. Planned evaluation will include assessment of discrimination, calibration, robustness, fairness across demographic groups, and clinical utility. Model updates will be governed through a version-controlled deployment framework to ensure reproducibility and traceability of algorithmic changes over time.

Future studies will also evaluate user trust, explainability, and the impact of AI-assisted decision support on referral pathways, workflow efficiency, and patient outcomes.

## 6. Prospective Evaluation Protocol

### 6.1 Study Design

The ASHA Assist India platform will undergo a prospective implementation study to evaluate its feasibility, usability, workflow integration, and operational performance within routine community healthcare settings. The study is designed as a phased implementation project, beginning with pilot deployment in selected rural communities before broader expansion.

The primary purpose of the pilot phase is not to evaluate the diagnostic accuracy of the proposed artificial intelligence modules but to assess whether the integrated digital platform can be successfully incorporated into routine workflows involving community health workers, primary healthcare providers, and referral hospitals. The evaluation will therefore focus on implementation outcomes, user experience, system performance, and operational feasibility.

### 6.2 Study Setting

The pilot implementation is planned within rural primary healthcare settings in India where Accredited Social Health Activists (ASHAs) routinely conduct community-based health visits. The platform is intended to operate across the existing healthcare hierarchy, including community households, Primary Health Centres (PHCs), and referral hospitals.

Site selection will consider:

- availability of ASHA workers,
- internet connectivity,
- existing referral pathways,
- willingness of healthcare facilities to participate,
- institutional approvals.

### 6.3 Study Population

The proposed evaluation will include four stakeholder groups.

#### Community Participants

Adults participating in community screening programmes who provide informed consent.

#### ASHA Workers

Accredited Social Health Activists involved in routine community healthcare delivery.

#### Primary Healthcare Professionals

Medical officers, nurses, and other PHC personnel responsible for reviewing participant records and coordinating referrals.

#### Referral Hospital Specialists at secondary and tertiary level of care

Clinicians involved in specialist evaluation and management following referral.

### 6.4 Inclusion Criteria

The planned study will include:

- adults aged 18 years or older,
- individuals willing to provide informed consent,
- residents within the selected implementation areas,
- ASHA workers participating in the pilot programme,
- participating PHCs and referral hospitals.

### 6.5 Exclusion Criteria

Participants may be excluded if they:

- decline informed consent,
- are unable to participate in follow-up,
- provide incomplete baseline information that precludes meaningful assessment,
- withdraw from the study at any stage.

### 6.6 Planned Data Collection

The platform is designed to collect multiple categories of information during routine community screening and follow-up activities.

#### Demographic Information

- Age
- Sex
- Education
- Occupation
- Family history
- Household characteristics

#### Lifestyle Factors

- Tobacco use
- Alcohol consumption
- Physical activity
- Dietary habits

#### Clinical Variables

- Blood pressure
- Blood sugar levels
- BMI
- Other NCD risk factors

#### Medication history

- Previous cardiovascular disease
- Follow-up Information
- Referral status
- Clinical outcomes
- Medication adherence
- Lifestyle modification
- Follow-up visits
- 6.7 Outcome Measures

#### Primary Outcomes

The pilot study will evaluate:

- System usability
- User acceptability
- Operational feasibility
- Workflow integration
- Secondary Outcomes

#### Secondary outcomes include

- Data completeness
- Referral completion rate
- Follow-up compliance
- Time required for participant screening
- User satisfaction
- System uptime
- Synchronization success
- Technical performance
- AI recommendation acceptance rate (future module)

### 6.8 Data Management

All participant information will be stored using encrypted communication channels and secure cloud infrastructure. Role-based authentication, audit logging, and controlled access permissions are planned to protect participant confidentiality. Personally identifiable information will be accessible only to authorized healthcare personnel according to predefined user roles.

Data backups, disaster recovery procedures, and routine security monitoring are planned to ensure platform reliability and data integrity throughout the implementation period.

### 6.9 Ethical Considerations

Prospective deployment will commence only after approval from the Institutional Ethics Committee (IEC) and relevant institutional authorities.

All participants will provide informed consent before enrolment. The study will comply with the Declaration of Helsinki, applicable national ethical guidelines, and relevant data protection regulations governing digital health research.

The platform will be ABDM and DPDP act complaint

## 7. Data Governance, Security, Privacy, and Interoperability

### 7.1 Data Governance Framework

The ASHA Assist India platform is designed to support secure and responsible management of health information throughout the data lifecycle. The proposed governance framework encompasses data collection, storage, transmission, access, analysis, archival, and eventual disposal, ensuring that participant information is handled in accordance with applicable ethical and regulatory standards.

Data ownership will remain with the participating institutions and study participants in accordance with institutional policies and national regulations. The platform is intended to maintain comprehensive audit trails documenting data creation, modification, and access to support transparency and accountability during prospective deployment.

### 7.2 Data Security

Security has been incorporated as a core design principle of the proposed platform. Multiple technical safeguards are planned to protect participant information against unauthorized access, modification, or disclosure.

The planned security framework includes:

- End-to-end encryption for data transmission.
- Encryption of data stored within backend databases.
- Secure user authentication.
- Role-based authorization.
- Session management.
- Automatic logout following inactivity.
- Audit logging of user activities.
- Secure cloud-based backup and disaster recovery.
- Regular security monitoring and system maintenance.

The platform is designed to ensure that users access only information necessary for their assigned responsibilities within the healthcare workflow.

### 7.3 Privacy Protection

Protection of participant privacy is fundamental to the proposed implementation. Personally identifiable information will be collected only when necessary for healthcare delivery and follow-up. Wherever possible, analytical datasets will utilize de-identified or pseudonymized records to minimize privacy risks during secondary analyses.

The platform is intended to support:

- Controlled access to identifiable information.
- Participant consent management.
- Data minimization.
- Secure storage of identifiable records.
- Separation of clinical identifiers from analytical datasets.
- Controlled data export for approved research activities.

These measures are designed to reduce the risk of inadvertent disclosure while preserving the utility of the data for clinical and research purposes.

### 7.4 Interoperability

Interoperability is an important consideration for long-term sustainability of digital health platforms. The proposed architecture is designed to facilitate communication with external health information systems through standardized application programming interfaces (APIs) and widely accepted interoperability standards.

Future integration may include:

- Electronic Health Records (EHRs).
- Hospital Information Systems (HIS).
- Laboratory Information Systems (LIS).
- Health Information Exchanges (HIEs).
- National Digital Health Ecosystem services.
- Ayushman Bharat Digital Mission (ABDM) components, where appropriate and subject to regulatory approval.

The modular architecture is intended to allow future interoperability without requiring major structural changes to the platform.

### 7.5 Ethical Considerations

Prospective deployment of the ASHA Assist India platform will commence only after approval by the appropriate Institutional Ethics Committee (IEC) and other relevant regulatory authorities. All participants will provide informed consent before enrolment.

The platform is intended to support ethical use of artificial intelligence through a human-in-the-loop decision-support framework, ensuring that AI-generated recommendations supplement rather than replace clinical judgement. Final decisions regarding referral, diagnosis, or treatment will remain the responsibility of qualified healthcare professionals.

### 7.6 Regulatory Considerations

The platform is intended to operate in accordance with applicable national policies governing digital health, health information management, and personal data protection. During future implementation, the project will align with institutional governance requirements and evolving national digital health initiatives, including interoperability frameworks where appropriate.

As healthcare regulations continue to evolve, periodic review of governance policies, security measures, and data management procedures will be incorporated into future platform maintenance.

## 8. Expected Impact and Future Directions

### 8.1 Expected Public Health Impact

Stroke prevention remains one of the most cost-effective strategies for reducing long-term disability and premature mortality. The ASHA Assist India platform is intended to strengthen community-based stroke prevention by integrating digital health technologies within the existing rural healthcare delivery system rather than creating parallel healthcare pathways. By supporting standardized data collection, longitudinal follow-up, and coordinated referral across multiple levels of care, the proposed platform has the potential to improve continuity of care and facilitate earlier identification of individuals at increased risk of stroke.

Integration of community health workers into digital screening programmes may also enhance population coverage while reducing the documentation burden associated with traditional paper-based workflows. If successfully implemented, the platform could support more efficient communication between communities, Primary Health Centres, and referral hospitals, thereby strengthening preventive healthcare delivery within resource-constrained settings.

### 8.2 Expected Scientific Contributions

Beyond its potential public health applications, the proposed platform is expected to generate valuable longitudinal health data that can support future epidemiological and implementation research. Prospective data collected through routine community screening may facilitate the development, calibration, and validation of predictive models using population-specific datasets, thereby improving the relevance of analytical tools for Indian populations.

The modular architecture also provides a flexible research environment in which additional analytical modules can be incorporated without requiring major modifications to the overall platform. Future investigations may evaluate disease-specific prediction models, digital biomarkers, personalized preventive interventions, and implementation strategies for AI-assisted community healthcare.

### 8.3 Potential for Health System Integration

The platform has been designed with interoperability as a guiding principle, allowing future integration with broader national digital health initiatives where appropriate. Although the present protocol does not include external system integration, the modular backend architecture is intended to facilitate communication with electronic health records, laboratory information systems, and national digital health services in future implementation phases.

Such integration could support continuity of care across healthcare settings while minimizing duplication of clinical documentation and improving availability of longitudinal health information for healthcare providers.

### 8.4 Scalability

The proposed architecture is designed to support incremental expansion beyond the initial pilot implementation. Following successful evaluation, future deployments may include additional Primary Health Centres, district hospitals, and geographically diverse populations. The modular design also allows adaptation of the platform to other non-communicable diseases, including hypertension, diabetes mellitus, cardiovascular disease, chronic kidney disease, and chronic respiratory disorders.

Furthermore, the overall framework may be adapted for use in other low- and middle-income countries employing community health worker programmes with similar healthcare structures.

### 8.5 Future Development of Artificial Intelligence Modules

The artificial intelligence framework described in this protocol represents an evolving research component rather than a completed clinical decision-support system. Future work will focus on the development and prospective validation of population-specific predictive models using longitudinal datasets collected through platform deployment.

Potential future developments include:

- Dynamic risk prediction using repeated longitudinal observations.
- Explainable artificial intelligence approaches to improve model transparency.
- Federated learning strategies enabling collaborative model development without centralized sharing of sensitive health information.
- Fairness assessment across demographic and socioeconomic groups.
- Continuous model updating through prospective learning while maintaining rigorous validation procedures.

Each analytical module will undergo independent validation before consideration for routine clinical use.

### 8.6 Dissemination Strategy

Findings arising from future implementation studies will be disseminated through peer-reviewed publications, conference presentations, technical reports, and engagement with public health stakeholders. Where appropriate, software components, technical documentation, and implementation resources may be made available under suitable open-source or institutional licensing arrangements to facilitate transparency, reproducibility, and future collaboration.

### 8.7 Long-Term Vision

The long-term vision of ASHA Assist India is to establish a scalable digital health ecosystem that supports preventive healthcare through integration of community health workers, structured digital workflows, and evidence-based clinical decision support. Rather than focusing solely on stroke prevention, the underlying platform is intended to provide a flexible foundation for future digital health programmes addressing multiple chronic diseases within primary healthcare settings.

Successful implementation and evaluation of the proposed protocol may contribute to broader understanding of how digital technologies can strengthen community health systems while preserving the central role of frontline healthcare workers in preventive care.

## 9. Conclusion

The ASHA Assist India project presents a protocol and technical design for an integrated digital health platform intended to support community-based stroke prevention through India’s Accredited Social Health Activist (ASHA) network. By combining structured digital workflows, interoperable system architecture, and planned artificial intelligence–assisted decision support within a unified platform, the proposed framework seeks to strengthen coordination between citizens, community health workers, primary healthcare centres, and referral hospitals.

Unlike many digital health initiatives that focus primarily on predictive algorithms, the proposed platform emphasizes the broader healthcare ecosystem, integrating data collection, longitudinal monitoring, referral management, and clinical decision support within routine community healthcare workflows. The modular architecture has been designed to support scalability, interoperability, and future expansion while maintaining flexibility for incorporation of additional disease-specific analytical modules and integration with national digital health initiatives.

The present manuscript describes the planned implementation strategy, system architecture, evaluation framework, and governance principles that will guide future development and prospective assessment of the platform. Clinical validation, implementation outcomes, and assessment of the artificial intelligence modules remain the subject of future studies following pilot deployment and appropriate ethical approvals.

If successfully implemented and evaluated, ASHA Assist India has the potential to provide a scalable framework for digitally enabled community health programmes supporting stroke prevention and other non-communicable diseases in resource-constrained settings. More broadly, the protocol aims to contribute to the growing field of implementation science by providing a transparent and reproducible roadmap for the development, deployment, and evaluation of AI-enabled digital health platforms within primary healthcare systems.

## Protocol Status Version: 1.0

**Current Stage:** Protocol and technical design completed.

**Software Development:** Planned.

**Internal Testing:** Planned.

**Pilot Deployment:** Pending Institutional Ethics Committee approval.

**Prospective Evaluation:** Not yet initiated.

**Clinical Validation:** Planned following pilot implementation.

**Artificial Intelligence Modules:** Conceptual framework described; model development and validation will be reported separately.

**Expected Timeline:** To be determined following institutional approvals.

## Data Availability

No new data was generated in this study

## References

1. GBD 2021 Stroke Collaborators. Global, regional, and national burden of stroke and its risk factors, 1990–2021. Lancet Neurol. 2024;23:973–1003.

2. Feigin VL, Brainin M, Norrving B, Martins SO, Sacco RL, Hacke W, et al. World Stroke Organization Global Stroke Fact Sheet 2025. Int J Stroke. 2025.

3. Pandian JD, Sudhan P. Stroke epidemiology and stroke care services in India. J Stroke. 2013;15(3):128–134.

4. Kamalakannan S, Gudlavalleti ASV, Gudlavalleti VSM, Goenka S, Kuper H. Incidence and prevalence of stroke in India: A systematic review. Indian J Med Res. 2017;146:175–185.

5. World Health Organization. Global Strategy on Digital Health 2020–2025. Geneva: World Health Organization; 2021.

